# In which journals do family and community physicians in Brazil publish? The *Trajetórias MFC* project

**DOI:** 10.1101/2020.06.24.20138537

**Authors:** Leonardo Ferreira Fontenelle, Miguel Henrique Moraes de Oliveira, Stephani Vogt Rossi, Diego José Brandão, Thiago Dias Sarti

## Abstract

**Introduction:** Authors choose scholarly journals not only to advance their careers but also to interact with their respective scholarly communities.

**Objective:** To describe the journals where family and community physicians in Brazil publish their work.

**Methods:** In late 2018, we compiled a nationwide list of family and community physicians, and downloaded their curricula vitae (CV) from the Lattes Platform. We extracted data on their complete journal articles from their CV, completed these data with queries to CrossRef, VHL/LILACS, and PubMed/MEDLINE, and obtained data on the journals with queries to the United States NLM Catalog.

**Results:** We found 3622 unique articles, published by 1014 journals. The most productive journal was RBMFC (*Revista Brasileira de Medicina de Família e Comunidade*), which published 353 (9.7%) of these articles. About one in six articles were published in journals on family practice or primary health care. The proportion of articles published in journals in Brazil decreased during the study period from 83.3% to 59.0%.

**Conclusion:** As in other countries, family and community physicians in Brazil usually publish in the national journal dedicated to their scholarly community, while also publishing extensively in journals from other disciplines. The increasing proportion of articles published in journals outside Brazil suggests primary care research in Brazil is increasingly of international relevance.

**Biography:** **Leonardo Ferreira Fontenelle:** Professor at Universidade Vila Velha. Graduated in medicine from Universidade Federal do Espírito Santo; medical residency in family and community medicine from Universidade de São Paulo Medical School of Ribeirão Preto; master’s degree in community health at Universidade de São Paulo Medical School of Ribeirão Preto; Ph.D. in epidemiology at Universidade Federal de Pelotas, Medical School.

URL (Lattes): http://lattes.cnpq.br/9234772336296638

Competing interests: see below.

Contribution: Conceptualization; Data curation; Investigation; Methodology; Project administration; Software; Supervision; Visualization; Writing – original draft.

**Miguel Henrique Moraes de Oliveira:** Medical doctor graduated from Universidade Vila Velha.

URL (Lattes): http://lattes.cnpq.br/0608163043063251

Competing interests: None.

Contribution: Data curation; Investigation; Writing – original draft.

**Stephani Vogt Rossi:** Medical student at Universidade Vila Velha.

URL (Lattes): http://lattes.cnpq.br/7614124748693457

Competing interests: None.

Contribution: Data curation; Writing – review & editing.

**Diego José Brandão:** Professor at Universidade Vila Velha. Graduated in medicine from Escola Superior de Ciências da Santa Casa de Misericória de Vitória (EMESCAM); medical residency in family and community medicine at Universidade de São Paulo Medical School; Ph.D. in preventive medicine at Universidade de São Paulo Medical School.

URL (Lattes): http://lattes.cnpq.br/5130371131757497

Competing interests: see below.

Contribution: Conceptualization; Methodology; Writing – review & editing.

**Thiago Dias Sarti:** Professor at Universidade Federal do Espírito Santo. Graduated in medicine from Escola Superior de Ciências da Santa Casa de Misericória de Vitória (EMESCAM); medical residency in family and community medicine at Universidade Estadual do Rio de Janeiro; master’s in collective health at Universidade Federal do Espírito Santo, Center of Health Sciences; Ph.D. in public health at Universidade de São Paulo, School of Publich Health.

URL (Lattes): http://lattes.cnpq.br/7489127535403969

Competing interests: see below.

Contribution: Conceptualization; Methodology; Writing – review & editing.

## Introduction

Scholarly journals are essential infrastructure for their respective scholarly disciplines and communities.^1–3^ While publishing in journals with high prestige or impact factor advances the careers of individual researchers, publishing in journals with the right readership communicates the scholarly output to the corresponding scholarly community, thus contributing to its intellectual conversations.^2–5^ Family medicine journals are thus strategic for strengthening the capacity for primary care research,^6–8^ which in turn informs primary care and reinforces its status as a scholarly discipline.^9–11^

The status of family medicine or primary care as scholarly disciplines varies substantially across countries.^7^ In Brazil, family and community medicine was recognized as a medical specialty in 1981,^12^ but most physicians in primary care are not certified family and community physicians.^13^ Neither family and community medicine, nor primary care is recognized as scholarly disciplines for research funding or *stricto sensu* postgraduate education (master’s and doctorates).

The lack of recognition of their scholarly discipline might compel family and community physicians in Brazil to engage in “intellectual discussions” (to borrow from Wakeling *et al*. ^3^) with neighboring scholarly communities, such as other medical specialties or public health, rather than among themselves. In this study, our objective was to describe the scholarly journals where they publish their scholarly output, as an approach to better understanding the state of primary care research in Brazil.

## Methods

This was an observational, exploratory study, using administrative data from multiple sources, as part of the *Trajetórias MFC* project.^14^ Ethical approval was obtained from the research ethics committee in Universidade Vila Velha (certificate 02957118.2.0000.5064), and data on specialist certification was obtained with permission from SBMFC (*Sociedade Brasileira de Medicina de Família e Comunidade*, the Brazilian Society on Family and Community Medicine). Three authors (LFF, DJB, and TDS) are family and community physicians and discussed the research project informally with their peers, but stakeholders were not formally included in the study planning.

### Data sources

As previously described,^14^ we compiled the list of family and community physicians in Brazil using data from SisCNRM (*Sistema da Comissão Nacional de Residência Médica*, the national information system on medical residency) in November 2018 and SBMFC in late December 2018, and obtained their curricula vitae from the Lattes Platform (the national information system on the evaluation of science, technology, and innovation ^15^), also in late December 2018.

The list of complete journal articles was obtained from the family and community physicians’ curricula vitae. During late 2019 through early 2020, we obtained missing DOI (digital object identifiers) from CrossRef using rcrossref 0.9.2,^16^ PubMed identifiers (PMID) from the United States’ National Library of Medicine (NLM) through rentrez 1.2.2,^17,18^ and LILACS identifiers from the Pan American Health Organization’s Virtual Health Library (VHL; in Portuguese: *Biblioteca Virtual em Saúde*, BVS) using httr 1.4.1 ^19^. We also obtained the corresponding journal titles and ISSN (International Standard Serial Number) from the curricula vitae and then used rentrez in July 2019 to find the corresponding journal records in the NLM Catalog, which is not restricted to journals indexed in PubMed/MEDLINE.^20^

Throughout the data acquisition process, all data were extensively verified by up to three authors (between LFF, MHMO and SVR). For example, we made sure every journal was always listed with the same name, consulting when necessary the corresponding journal entries in the NLM Catalog, the ISSN International Center ROAD (Directory of Open Access scholarly Resources)^21^ and the VHL Serials in Health Sciences (SeCS; in Portuguese, *Seriados em Ciências da Saúde*). We also made sure each journal article had the same LILACS identifier, PMID, and/or DOI when listed more than once, and furthermore considered journal, volume and starting page to identify articles published by more than one family and community physician.

We defaulted to trusting data contained in the curricula vitae. For example, we verified the DOI retrieved from CrossRef but not the ones provided in the curricula vitae. Sometimes, however, verifying the internal consistency of programmatically acquired data revealed mistakes in the curricula vitae requiring data editing (for example, wrong page number) or even record removal (for example, journal articles entered more than once by the same author, poster abstracts published in supplements of scholarly journals, or articles published in clearly non-scholarly journals).

### Variables and analysis

The scholarly output of family and community physicians from Brazil was described using the year of publication and the journals’ titles, country, and major subjects. We made these data freely available at Zenodo.^22^

Journal articles were included only if they were published during or after the year of specialization, which was taken to be the year of certification by SBMFC or conclusion of the medical residency, whichever came first. The year of publication was categorized in five-year periods, except for the first period, which included the years 1985 (first included article) through 1998 (last year before the Lattes Platform became online).

The MeSH terms for the journal’s major subjects were derived from the corresponding entries from the NLM Catalog. Because the number of major subjects per journal ranged from zero to six, we weighted major subjects by the inverse of their number within a journal. For the sake of simplicity, we did not weight the journal articles by the number or proportion of family and community physicians among the authors.

The description of the most productive (that is, with more included articles) journal titles emphasized those amounting to one-third of the included articles.

## Results

From the 6238 family and community physicians in Brazil as of late December 2018, 4065 (65.2%) had a curriculum vitae in the Lattes Platform. In total, 1115 (17.9%) had published one or more journal articles, with an average of 5.0 articles per author (median 2, interquartile range [IQR] 1–4). Considering only articles published during or after the year of specialization, 637 (10.2%) of the family and community physicians had published an average of 5.7 articles per author (median 2, IQR 1–5).

Some journal articles appeared in our data more than once, because they had been written in collaboration by more than one family and community physician. After removing these duplicates, there were 3622 unique journal articles. Publication years ranged from 1985 to 2018, with a median of 2012 and an IQR of 2008–2016.

These 3622 unique journal articles were published by 1014 unique scholarly journals. The 14 most productive journals published 1231 (34.0%) articles (Table 1), while the 847 least productive journals published 1144 (31.6%) articles by family and community physicians in Brazil. *Revista Brasileira de Medicina de Família e Comunidade* (RBMFC) was the most productive journal since its inception in 2004: it published 353 (9.7%) articles, more than the next three journals combined.

**Table 1.**
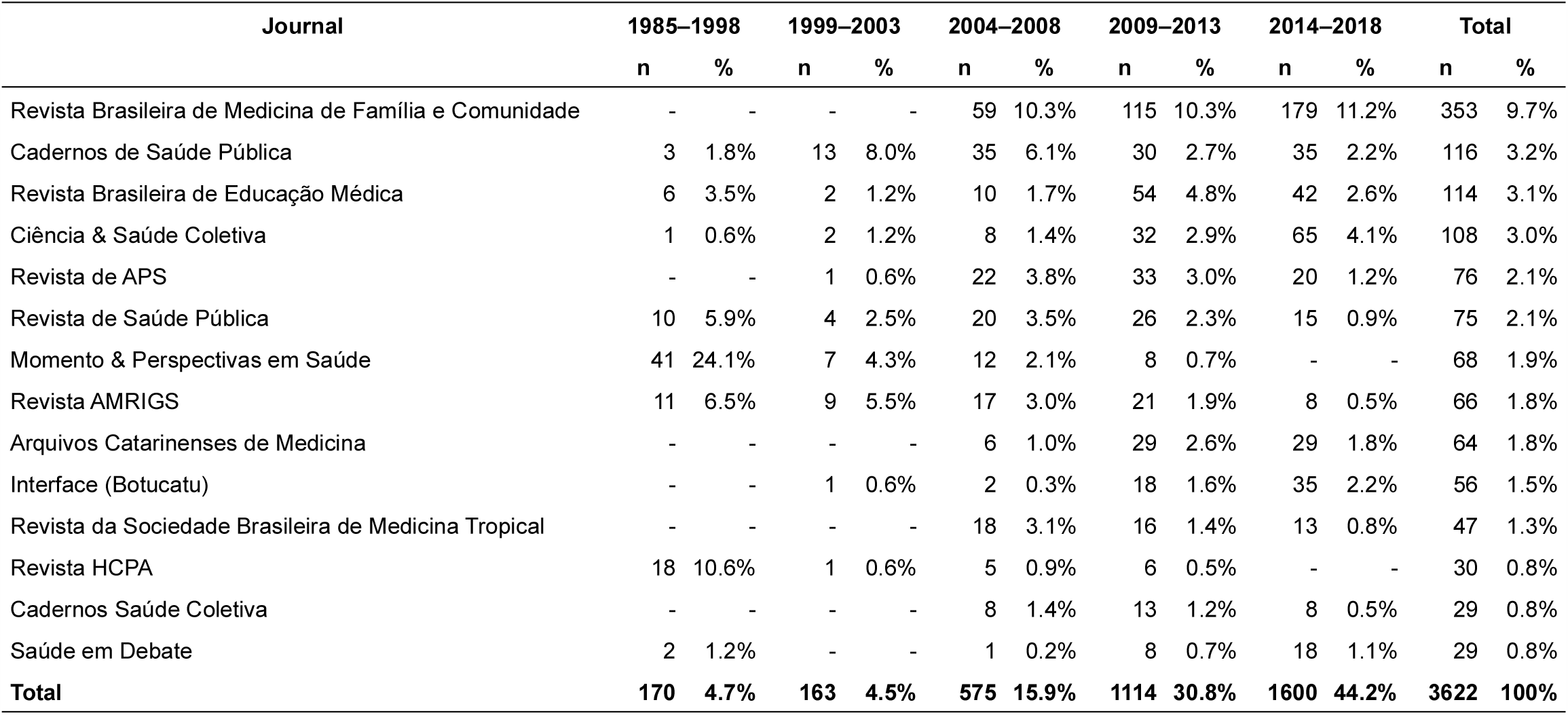
Scholarly journals publishing most articles by family and community physicians in Brazil, up to December 2018.

The NLM Catalog included 735 (72.5%) of the scholarly journals, accounting for 3091 (85.3%) of the journal articles. Medicine in general was the most frequent major subject, with general medicine journals having published 479.0 (16.6%) articles by family and community physicians in Brazil (Table 2). Surprisingly, the most productive journal in this category was *Revista de APS*, which is cataloged as having medicine as its only major subject, rather than primary health care as expected. The next journals in this category were, in order: *Revista AMRIGS, Arquivos Catarinenses de Medicina*, and PLoS One.

**Table 2.** Most frequent major subjects of scholarly journals publishing articles by family and community in Brazil, up to 2018.

| Journal | 1985–1998 |  | 1999–2003 |  | 2004–2008 |  | 2009–2013 |  | 2014–2018 |  | Total |  |
| --- | --- | --- | --- | --- | --- | --- | --- | --- | --- | --- | --- | --- |
|  | n | % | n | % | n | % | n | % | n | % | n | % |
| Medicine | 25.3 | 17.1% | 26.0 | 22.2% | 78.3 | 16.3% | 171.0 | 18.4% | 178.3 | 14.7% | 479.0 | 16.6% |
| without Revista de APS | 25.3 | 17.1% | 25.0 | 21.4% | 56.3 | 11.7% | 138.0 | 14.8% | 158.3 | 13.0% | 403.0 | 13.9% |
| Public Health | 15.0 | 10.1% | 22.0 | 18.8% | 88.0 | 18.3% | 135.0 | 14.5% | 185.4 | 15.2% | 445.4 | 15.4% |
| Primary Health Care | - | - | - | - | 59.0 | 12.2% | 116.0 | 12.5% | 183.3 | 15.1% | 358.3 | 12.4% |
| with Revista de APS | - | - | 1.0 | 0.9% | 81.0 | 16.8% | 149.0 | 16.0% | 203.3 | 16.7% | 434.3 | 15.0% |

The second most frequent major subject was public health, with 445.4 (15.4%) articles (Table 2). The most productive journals in the group were *Cadernos de Saúde Pública* and *Ciência & Saúde Coletiva*, followed by *Revista de Saúde Pública* and *Cadernos Saúde Coletiva*.

The third most frequent major subject was primary health care, with 358.3 (12.4%) articles (Table 2). Almost all articles in this group were published in RBMFC, which is cataloged with primary health care as its major subject and family practice as its secondary one. If *Revista de APS* were included in this group, primary health care would become the second most frequent major subject, ahead of medicine and close to public health.

Combined, medical education and health education accounted for 200.5 (3.5%) articles. Within each major subject, most articles were published by *Revista Brasileira de Educação Médica* and *Interface (Botucatu)*, respectively.

Family practice was the ninth most frequent major subject, with 38.2 (1.3%) articles. The main journals were the Annals of Family Medicine and Family Practice (data not shown).

The NLM Catalog-listed journals had their headquarters in 42 different countries. Journals from Brazil published 2054 (66.5%) of the articles, being followed by journals from England (363, 11.8%) and the United States (336, 10.9%). Over time, the proportion of articles published in journals in Brazil dropped from 83.3% to 59.0% (Figure 1), while the proportion in journals from other countries rose accordingly.

**Figure 1.**
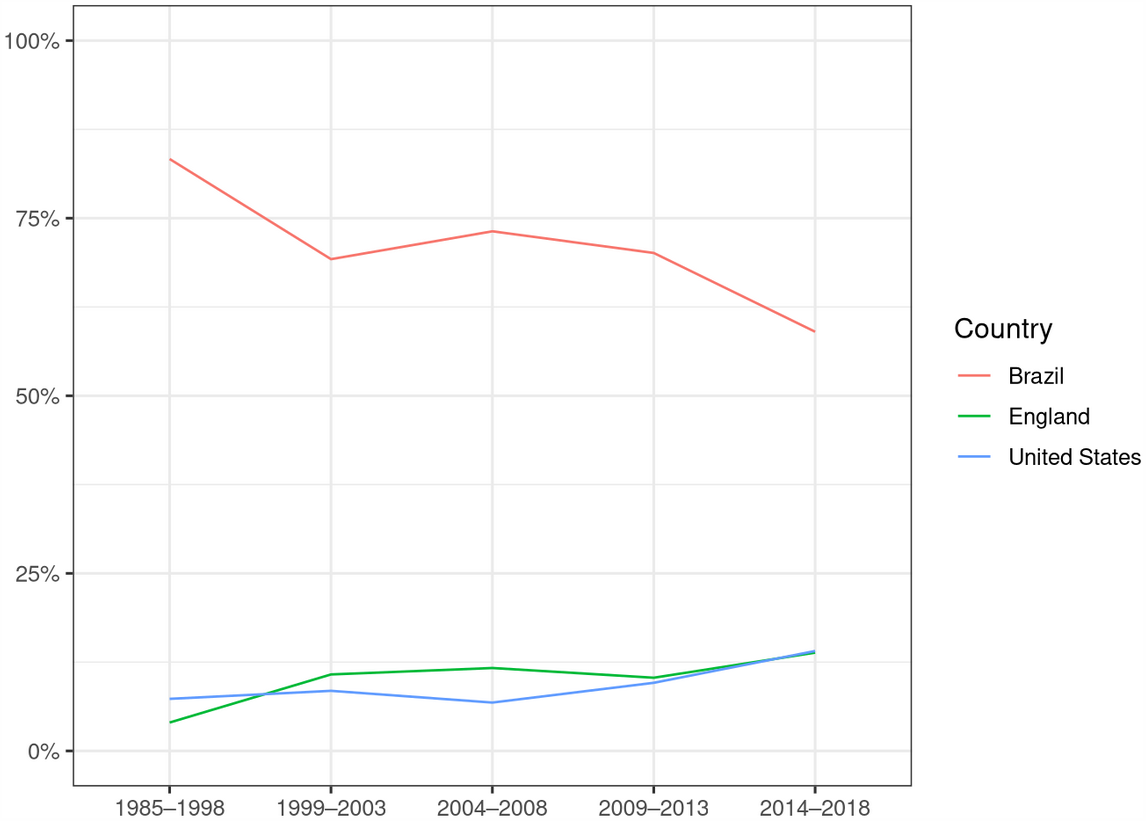
Time trend of publication countries of articles by family and community physicians in Brazil, up to 2018.

## Discussion

As of late December 2018, family and community physicians in Brazil had published their scholarly articles in more than a thousand journals, from tens of countries. Nevertheless, their national disciplinary journal was also the most prolific one, publishing one in ten of their articles. Because traditional journal-level metrics have not favored RBMFC, we can infer family and community physicians value the opportunity to engage in intellectual conversations with their own scholarly community. Such preference for national journals dedicated to their scholarly discipline has also been found among family physicians from the United States,^23^ faculty in university departments of family medicine in the United States,^24^ and primary care researchers from Spain.^25,26^ This pattern seems to confirm the importance of national and regional organizations of family medicine maintaining journals to strengthen the scholarly status and research capacity of the discipline.^6–8^

On the other hand, only one in six articles were published in journals about family practice or primary health care (with or without *Revista de APS*); the other were scattered in journals about public health, medicine in general, medical/health education and a myriad of other subjects. This diversity of journal subjects is well documented for family physicians and for faculty in departments of family medicine, at least in the United States.^23,24^ Being at the border of clinical medicine with applied social sciences,^27,28^ family and community medicine has ample opportunity for interdisciplinary research, which might or might not be published in journals about family practice or primary health care. Furthermore, journals of bordering disciplines may be more attractive for career progression because of publishing more basic research and thus having higher journal-level citation metrics.^29^

Although interdisciplinary publication is an international characteristic of family medicine as a scholarly discipline, the Brazilian emphasis on public health deserves further scrutiny. Of the 14 journals accounting for a third of the articles written by family and community physicians from Brazil, six are public health journals per the NLM Catalog. One reason might be that these are actually journals about collective health, which in Latin America is a scholarly discipline comprising public health, epidemiology, and human and social sciences in health,^30^ among other definitions.^31^ This wider scope (compared to public health) means collective health frequently includes health research which is not strictly biomedical but would be considered medical research in other contexts.

Another reason might be that most family and community physicians in Brazil earn their master’s and Ph.D. degrees in collective health, not medicine.^14^ This prevalence of postgraduate degrees in collective health might explain why, as reported by Almeida et al.,^28^ primary care research in Brazil has a focus on healthcare services and policies, instead of clinical research. This focus, in turn, might explain why family and community physicians submit so much research to public health (collective health) journals.

Overall, one-third of the articles were published in journals in other countries, and this proportion seems to be increasing over time. In other words, at least one-third of the scholarly output of family and community physicians in Brazil is of international relevance: otherwise, there would be little reason for publishing abroad. We believe the proportion of internationally relevant articles is even greater, because journals from Brazil may be of international relevance themselves, and individual articles might be internationally relevant even if published in national journals. Moreover, the increasing proportion of articles published in journals in other countries suggests primary care research in Brazil is increasingly of internal relevance.

Our findings should be interpreted with some limitations in mind, despite deriving from carefully curated data obtained from authoritative sources. For instance, consistently with Brazilian norms on medical specialization, we included only physicians concluding the medical residency since 1981, as well as those undergoing special certification since 2004. When family and community medicine (then “general community medicine”) was recognized as a specialty, medical doctors having already concluded their medical residencies, or having experience working as family and community physicians, could not opt to be recognized as such. We can only wonder what scholarly communities do these unacknowledged family and community physicians interact with.

Furthermore, our study was restricted to the scholarly output published in peer-reviewed journals, because of their centrality to the advancement of scholarly disciplines and relative tractability to quantitative analysis. In consequence, our findings might not apply to scholarly communication through other media, such as conferences and textbooks. Furthermore, journal articles are not a direct measurement of knowledge advancement, because not all journal articles contribute equally.

Finally, the temporal context of our data should be considered. First, journal articles were eligible only if published after specialization, to ensure the relevance of the included articles to family and community medicine. The downside is that some of the excluded articles might be relevant, too. Last, our findings are probably overrepresentative of early career researchers, because most family and community physicians concluded their medical residencies or were certificated in the last 10 years ^14^ and, as a result, the specialty has a fairly young demographic profile.^32^

The young demographic profile of family and community medicine may constitute an opportunity for its development as an academic discipline. Primary care research has a wide spectrum, and research projects should be adequately distributed along this spectrum for primary care research to fulfill its role in the improvement of primary health care. If the sparsity of clinical research in primary care can be attributed to the *stricto sensu* postgraduate education received by family and community physicians in Brazil, the recently proposed master’s degree in family and community medicine ^33^ might be pivotal in improving clinical practice in primary health care.

## Data Availability

The authors made these data freely available at Zenodo.

https://doi.org/10.5281/ZENODO.3905255

## Acknowledgments

MHMO was supported by a scientific initiation scholarship from Universidade Vila Velha.

